# HEART rate variability biofeedback breathing programme for LOng Covid dysautonomia (HEARTLOC): results of a feasibility study

**DOI:** 10.1101/2023.09.09.23295208

**Authors:** J Corrado, N Iftekhar, SJ Halpin, M Li, R Tarrant, J Grimaldi, AD Simms, RJ O’Connor, AJ Casson, M Sivan

## Abstract

**Introduction:** Post-covid-19 syndrome, or Long covid (LC) refers to symptoms persisting 12 weeks after Covid-19 infection. LC comprises a wide range of dysautonomia symptoms, including fatigue, breathlessness, palpitations, dizziness, pain and brain fog. This study tested the feasibility and estimated the efficacy, of a Heart Rate Variability Biofeedback (HRV-B) technique via a standardised slow diaphragmatic breathing programme in individuals with LC.

**Methods and Analysis:** LC patients underwent a 4-week HRV-B intervention for 10 minutes twice daily for a total of 4 weeks using the Polar H10 ECG (Electrocardiogram) chest strap and Elite HRV phone application. Outcome measures C19-YRSm (Yorkshire Rehabilitation Scale modified), EQ5D-5L (EuroQol 5 Dimensions), Composite Autonomic Symptom Score (COMPASS-31), WHO Disability Assessment Schedule (WHODAS), and Root Mean Square of Successive Differences between heartbeats (RMSSD) using a Fitbit device were completed before and after the intervention. The study was pre-registered at clinicaltrials.gov NCT05228665.

**Results:** 13 participants (54% female, 46% male) completed the study with high levels of data completeness and adherence. There was a statistically significant improvement in C19YRS-m (p=0.001), EQ5D Global Health Score (p=0.009), COMPASS-31 (p=0.007), RMSSD (p=0.047) and EHODAS (p=0.02). Qualitative feedback suggested participants were able to use it independently, were satisfied with the intervention, and reported beneficial effects from the intervention.

**Conclusion:** This is the first study in the literature to report that HRV-B is a feasible intervention for LC and seems to be potentially improving symptoms of LC and dysautonomia.

**Article summary:** *Strengths and limitations of the study:*

- To our knowledge, this is the first study of HRV-B in long covid and dysautonomia and has shown the feasibility of a novel technology-based intervention in a home setting.
- There was a statistically significant improvement in LC symptoms, functional ability, quality of life and dysautonomia scores.
- The study provides an estimation of efficacy which will determine the sample size for a larger controlled trial in LC and dysautonomia.
- The main limitation of this study is the small sample size and uncontrolled design which might not give us an accurate estimate of efficacy.
- The sample was predominantly Caucasian participants, 54% of whom were female.
- The take-up for HRV-B in those with a lack of experience in using digital technology and those from less privileged backgrounds is unknown.

## Introduction

Post-covid-19 syndrome or Long covid (LC) refers to persistent symptoms 12 weeks after SARS-COV2 infection and includes symptoms of physical fatigue, cognitive fatigue or “brain fog”, breathlessness, pain and psychological distress.^1 2^ An estimated 1.9 million people are reported to be affected by LC in the UK alone.^3^ A GP at an average sized practice in the UK can expect to have 65 patients with Long covid.^4^ The condition can be highly debilitating for some, particularly middle-aged individuals who were previously functioning at a high level and in demanding vocational roles.^5^ Many will experience significant disruption to employment, social and caregiving roles, and participation in society.

Many LC symptoms such as palpitations, dizziness, fatigue, pain and breathlessness can be explained by dysfunction of the Autonomic Nervous System (ANS) or dysautonomia.^6-10^ Estimates of prevalence of dysautonomia in LC range from 2.5 to 67%.^8 10 11^ Usually the ANS can maintain a finely tuned state of homeostasis or mount an appropriate stress response where necessary.^10^ However, in dysautonomia, there is episodic dysregulation in the ANS, typically with sympathetic overdrive. Dysautonomia also plays a significant role in the symptomology of many long-term conditions including multiple sclerosis, Parkinson’s disease, diabetes mellitus, fibromyalgia, chronic fatigue syndrome and migraine.^12^

One way of estimating and measuring autonomic function is through heart rate variability (HRV), as cardiac rate and rhythm are controlled largely by the ANS. A low HRV is associated with sympathetic nervous system activation, also described as a state of ‘fight or flight’. Higher HRV corresponds with parasympathetic nervous system activation and is believed to reflect a state of rest and recovery. Lower HRV has been observed to be associated with fatigue and pain symptoms of chronic fatigue syndrome/myalgic encephalomyelitis (ME/CFS) and fibromyalgia ^13-15^, as well as other chronic physical and mental health pathologies including asthma, anxiety and stress.^13-17^

When physiological parameters such as HRV are monitored in real-time with self-regulation techniques such as breathing techniques to increase parasympathetic activity through vagus nerve activation to influence the parameters, this is known as biofeedback.^18 19^ To the best of our knowledge, there have not yet been any studies of HRV-B in LC. However, HRV-B using breathing techniques has been tested in other chronic conditions such as asthma^16^, depression ^20^, fibromyalgia^15^ and post-traumatic stress^21^ and a 2021 systematic review summarises these comprehensively. The optimal breathing frequency to produce maximal increase in HRV varies for each individual but on average is between 5.5 and 6 breaths per minute and is known as resonant breathing.^16 22 23^ Resonant breathing helps to restore autonomic balance due to increased baroreflex gain and vagal activation.^16 22-24^

The aim of this study is to determine the feasibility and impact of a structured HRV-B regime incorporating diaphragmatic breathing exercise, on LC and dysautonomia symptoms.

### Aims and objectives

The aim of this study was:

To assess the feasibility of a 4-week HRV-B structured breathing programme in individuals with LC.

The objectives include:

1. Do breathing exercises through HRV-B increase HRV amongst participants with LC?
2. Are consumer grade monitors appropriate technology to use for HRV-B in the domiciliary setting?
3. Does regular HRV-B have any effect on LC and dysautonomia symptoms?

## Methods

### Study design

This was a phase 2 uncontrolled open-label feasibility study of a home technology-based HRV-B in 15 individuals with LC. Participants were identified through the Leeds covid-19 Rehabilitation Service, based at Leeds Community Healthcare NHS Trust. The total study period was 6 weeks for each participant. The inclusion criteria were: age ≥ 18 years, confirmed LC diagnosis as per the NICE criteria for post-covid syndrome ^1^, self-rating of at least ‘moderate’ or ‘severe’ on dysautonomia questions of palpitations or dizziness on the C19-YRSm ^25^; and abnormal NASA Lean Test (NLT)^26-28^ (HR increase of 30bpm or ≥120bpm, OR, BP decrease of 20mmHg systolic or 10 mmHg diastolic in the first 3 minutes of standing).

NLT is an accepted measure of cardiovascular instability and is conducted at initial assessment clinic for all LC service users in the Leeds covid rehabilitation service. The patient lies down for 2 to 5 minutes prior to the test with HR and BP taken each minute to calculate average supine values. They then stand with heels 6 inches from a wall and lean back against it with HR and BP taken each minute for 10 min. Abnormal results (as described above) are demonstrated through orthostatic hypotension or tachycardia on standing which are hallmarks of dysautonomia and therefore objectively quantifiable.

Exclusion criteria were inability to use the wearable or smartphone app technology, cognitive difficulties or mental health disorders causing inability to consent, cardiac arrhythmia, unstable respiratory disease (except asthma management)

### Equipment and intervention

To collect medium-term HRV data, participants wore a Fitbit Charge 5 smartwatch for a total of 6 weeks. The HRV-B element was conducted using a Polar H10 chest strap for 10 minutes twice daily for a total of 4 weeks (Figure 1). This connected via Bluetooth to the Elite HRV smartphone app which was downloaded to participants’ phones. Participants aimed to increase their HRV score as displayed in Elite HRV in real time using a diaphragmatic breathing technique (as explained in the protocol paper ^29^). Omron M2 blood pressure monitor (endorsed by the British Hypertension Society) was used to conduct NLT in clinic and the adapted Autonomic Profile (aAP)^30^.

**Fig 1.**
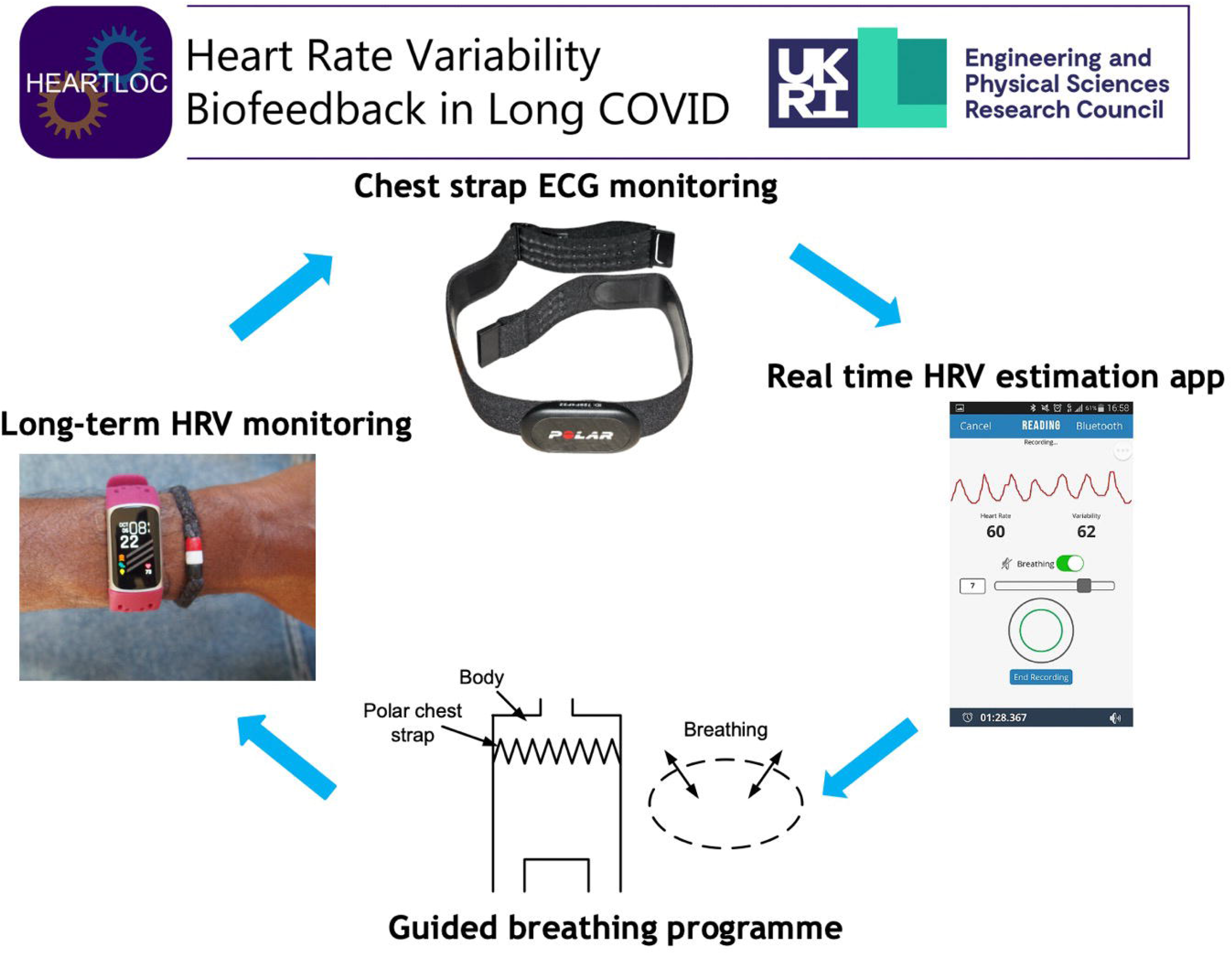
Heart Rate Variability Biofeedback (HRV-B) using a breathing technique and chest strap for real time HRV monitoring. Polar H10 picture from Wikimedia commons, reprinted under CC BY-SA 3.0 license. EliteHRV screenshot from Wikimedia commons, reprinted under CC BYSA 4.0 license.

### Insert Fig 1 about here

Participants were issued with a Fitbit Charge 5 device and smartphone application. They wore the Fitbit all the time during the 6 week period. This device collected nightly HRV data along with other measures of sleep.

They also received a Polar H10 chest strap which was connected to the Elite HRV smartphone app for the HRV-B element of the intervention. This started after an initial 1 week baseline period in which HRV data was collected only by Fitbit to allow a baseline of HRV data for comparison to post-intervention. The breathing intervention lasted for 10 minutes twice daily for a total of 6 weeks.

### Measures

**The primary outcome measure** was the C19YRSm, a self-reported patient-reported outcome measure to assess LC symptom severity, functional disability, and overall health status.

#### Secondary outcome measures

##### Fitbit Data measures

These included root mean square of successive differences between heartbeats (RMSSD), sleeping resting heart rate, non-rapid eye movement sleep heart rate (NREM HR), overall sleep score, composition score, revitalisation score, sleep duration score, deep sleep score

##### Patient Reported Outcome Measures

- Composite Autonomic Symptom Score (COMPASS 31)
- World Health Organisation Disability Assessment Schedule (WHODAS)
- EQ5D health related quality of life assessment (EQ-5D-5L)
- NASA Lean Test (NLT) – heart rate and blood pressure data
- adapted Autonomic Profile (aAP) – heart rate and blood pressure data

A summary of the schedule for the completion of outcome measures is shown in Table 1.

**Table 1.**
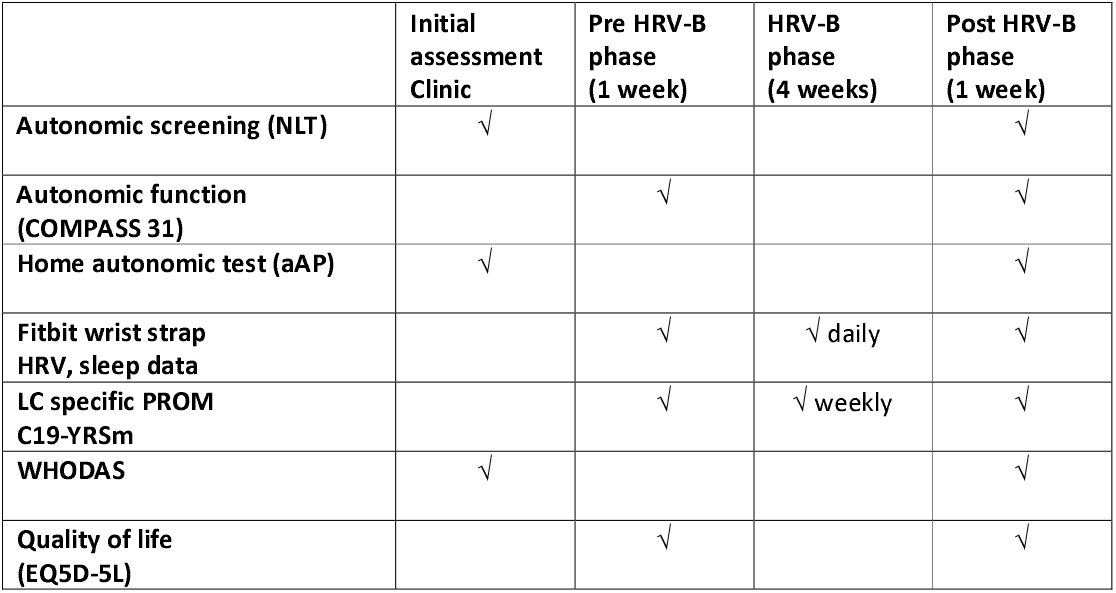
Outcome measures summary schedule.

|  | Initial assessment<br>Clinic | Pre HRV-B<br>phase<br>(1 week) | HRV-B<br>phase<br>(4 weeks) | Post HRV-B<br>phase<br>(1 week) |
| --- | --- | --- | --- | --- |
| Autonomic screening (NLT) | √ |  |  | √ |
| Autonomic function<br>(COMPASS 31) |  | √ |  | √ |
| Home autonomic test (aAP) | √ |  |  | √ |
| Fitbit wrist strap<br>HRV, sleep data |  | √ | √ daily | √ |
| LC specific PROM<br>C19-YRSm |  | √ | √ weekly | √ |
| WHODAS | √ |  |  | √ |
| Quality of life<br>(EQ5D-5L) |  | √ |  | √ |

### Analysis

Data from questionnaires were scored using standardised procedures for each measure. Data from the Fitbit was downloaded securely, cleaned and analysed using Python statistical platform. 2 participants were excluded from analysis, therefore a total of 13 individual datasets were analysed. Participant 6 was not able to complete the full 6 week study period due to unrelated health issues. Participant 12 was unwell during the final assessment with an acute respiratory infection. This was a cause extraneous to the study process and encompassed by our exclusion criteria therefore this participant was removed from data analysis. To explore the impact of the intervention, pre and post measures were compared on a within-participant basis using Wilcoxon signed rank tests. The pre-HRV phase was the baseline pre intervention period. The post-HRV-B phase data was used for post-intervention effect. Fitbit data and Elite HRV data for HRV was analysed during the HRV-B intervention phase and compared to pre-HRV-B data .A formal sample size calculation was not required for this feasibility study as it did not mimic a definitive randomised trial and aim was not to measure effect size.^31^

## Results

15 participants were enrolled in the study, out of which 13 completed the study and their data was included for the final analysis [Table 2]. Table 3 shows the results of the NLT for the 13 participants. There was a 92% compliance rate with the breathing intervention as a total of 720 sessions were completed over the 4-week period.

**Table 2.** Participant demographics.

| No | Age range | Sex | Ethnicity | LC Duration (months) | Comorbidities | Medication | Predominant LC symptoms | Study completion |
| --- | --- | --- | --- | --- | --- | --- | --- | --- |
| 1 | 40-50 | M | White | 13 | Anxiety<br>Depression | Amitriptyline<br>Codeine<br>Sumatriptan<br>Loratadine | Fatigue | Completed |
| 2 | 50-60 | F | White | 15 | Osteoarthritis | HRT<br>Naproxen | Breathlessness<br>Fatigue<br>Headache | Completed |
| 3 | 40-50 | M | White | 10 | Gastroesophageal reflux | Omeprazole | Fatigue<br>Breathlessness | Completed |
| 4 | 40-50 | F | Mixed | 20 | Adenomyosis<br>Hayfever |  | Pain<br>Fatigue<br>Breathlessness | Completed |
| 5 | 50-60 | M | White | 14 | Gastroesophageal reflux<br>Asthma<br>Skin disorder | Lansoprazole<br>Inhalers<br>Limocycline | Breathlessness | Completed |
| 6 | 60-70 | F | White | 12 | Osteoarthritis | Nil | Fatigue | Withdrew |
| 7 | 20-30 | F | White | 7 | Hayfever | Bisoprolol | Breathlessness<br>Pain | Completed |
| 8 | 30-40 | F | White | 25 | Anxiety/depression | Citalopram | Fatigue<br>Breathlessness<br>Sleep disturbance | Completed |
| 9 | 30-40 | M | White | 25 | Anxiety/depression | Citalopram | Fatigue | Completed |
| 10 | 40-50 | M | White<br>British | 15 | Nil | Nil | Fatigue | Completed |
| 11 | 10-20 | F | White | 12 | Nil | Oral contraception | Fatigue<br>Nausea<br>Breathlessness | Completed |
| 12 | 20-30 | F | White | 20 | Nil | Amitriptyline | Fatigue | Excluded |
| 13 | 40-50 | F |  | 11 | Crohn's disease<br>Migraine<br>ADHD | Azathioprine<br>Amitriptyline<br>Lisdexamphetamine<br>Ivabradine | Palpitations<br>Breathlessness | Completed |
| 14 | 40-50 | M | White | 17 | Asthma | Inhalers | Altered taste<br>Fatigue<br>Altered smell | Completed |
| 15 | 60-70 | F | White | 7 | Hypothyroidism<br>Hodgkins<br>Osteoporosis | Ivabradine<br>Thyroxine<br>Anastrozole<br>Alendronic acid<br>Aspirin | Fatigue<br>Breathlessness<br>Cough | Completed |

**Table 3.**
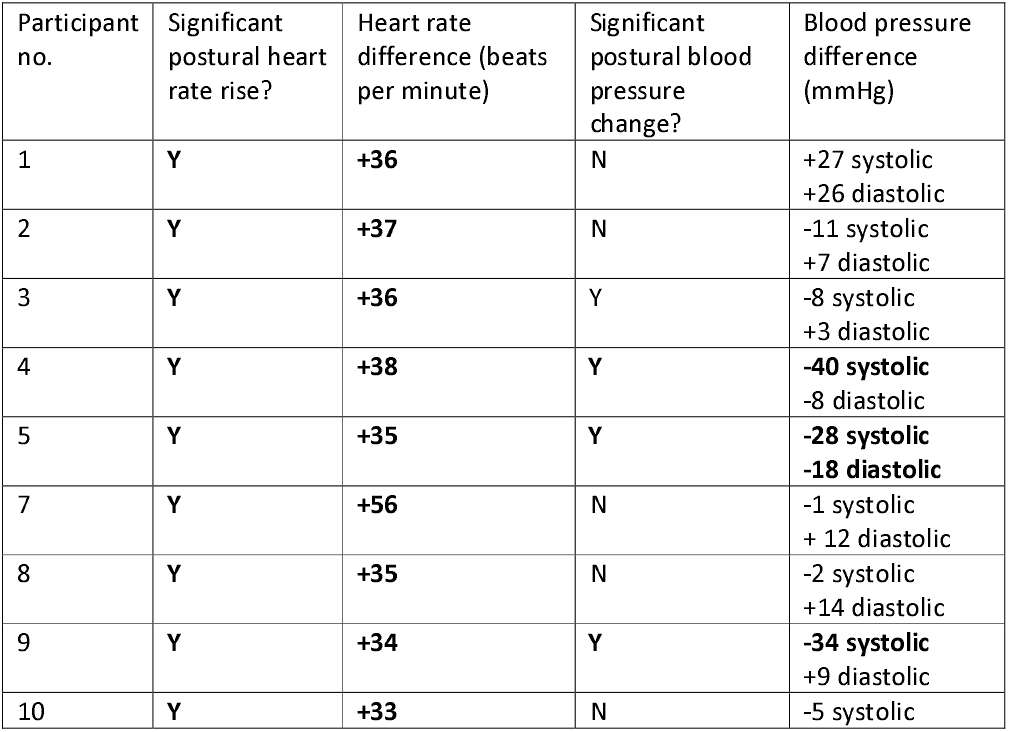
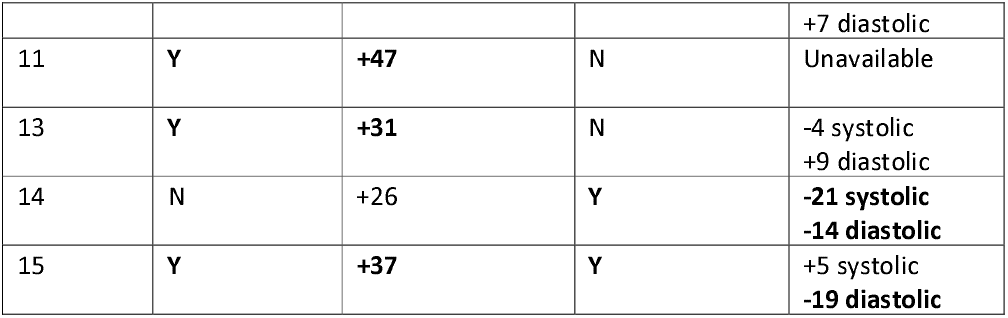
Results of baseline NASA Lean test.

**Table 4.** Change and effect size for the patient-reported outcome measure scores.

|  | Mean (SD) at baseline pre HRV-B phase | Mean SD at post HRV-B phase | Change (compared to baseline) | P value for paired pre/port difference | Effect Size (Cohen’s d) |
| --- | --- | --- | --- | --- | --- |
| **C19YRS-m symptom severity** | 19.54(4.37) | 13.46(6.54) | -5 | 0.0010 | 1.09316 |
| **C19YRS-m functional ability** | 7.615(3.885) | 5.231(4.902) | -2 | 0.0015 | 0.539022 |
| **C19YRS-m global health score** | 3.923(1.553) | 5.007(1.801) | +1 | 0.0137 | 0.644633 |
| **C19YRS-m total “other” LC symptoms** | 6.462(4.502) | 4.000(4.690) | -2 | 0.0059 | 0.535571 |
| **COMPASS-31** | 38.55(18.10) | 32.49(19.59) | -7.6 | 0.0078 | 0.047074 |
| **EQ5D-5L global health score** | 46.31(14.07) | 56.69(19.86) | +10 | 0.0083 | 0.007391 |
| **WHODAS disability scale** | 36.26(18.41) | 29.38(20.22) | -6.9 | 0.0215 | 2.029452 |

*C19YRS Symptoms Severity – range from 0-30, higher score = greater severity*
*C19YRS functional disability – range from 0-15, higher score = greater disability*
*C19YRS Total ‘other’ symptoms – total of 40 other miscellaneous symptoms possible*
*C19YRS global health score – range from 0-10-, higher score = greater total health perception*
*COMPASS-31 – range from 1 – 100, greater score = higher autonomic symptom burden*
*EQ5D-5L global health score – greater score = higher global health perception*

Statistically significant improvements were noted in Long Covid specific outcome measure C19YRS-m for symptom severity, functional ability, overall health score as well as total “other” symptoms, COMPASS31, and EQ5D-5L. The effect size for C19-YRSm was large.

There was a significant difference in RMSSD, a HRV measure reflecting parasympathetic activity^13^, between preHRV-B and post HRV-B phases (Table 5).

**Table 5.** HRV data from Fitbit device.

|  | Mean (SD) at baseline pre HRV-B phase | Mean SD at post HRV-B phase | P value for paired difference between post HRV-B and baseline | Effect Size (Cohen's d) |
| --- | --- | --- | --- | --- |
| Fitbit RMSSD Summaries | 34.19 (19.65) | 40.94 (29.36) | 0.0479* | 0.270202 |
| Fitbit Sleeping Resting HR | 67.84 (6.555) | 67.70 (5.932) | 0.8984 | 0.022395 |
| Fitbit NREM HR | 66.59 (7.606) | 66.15 (7.124) | 0.5417 | 0.05971 |
| Fitbit Overall Sleep Score | 76.78 (4.756) | 76.32 (5.083) | 0.8477 | 0.093454 |
| Fitbit Composition Score | 20.08 (1.288) | 19.70 (1.050) | 0.4143 | 0.323393 |
| Fitbit Revitalisation Score | 19.16 (1.878) | 18.75 (2.559) | 0.3394 | 0.182671 |
| Fitbit Sleep Duration Score | 37.53 (3.266) | 37.87 (2.932) | 0.6848 | 0.109554 |
| Fitbit Deep Sleep Score | 72.10 (22.14) | 69.20 (16.64) | 0.1909 | 0.14808 |

*Root Mean Square of Successive Differences between heartbeats (RMSSD)*
*Non-Rapid Eye Movement Heart Rate (NREM HR)*

Participants were asked how they felt about the technology and its effect on their LC symptoms. Some of the patient quotes are summarised in Table 6.

**Table 6.**
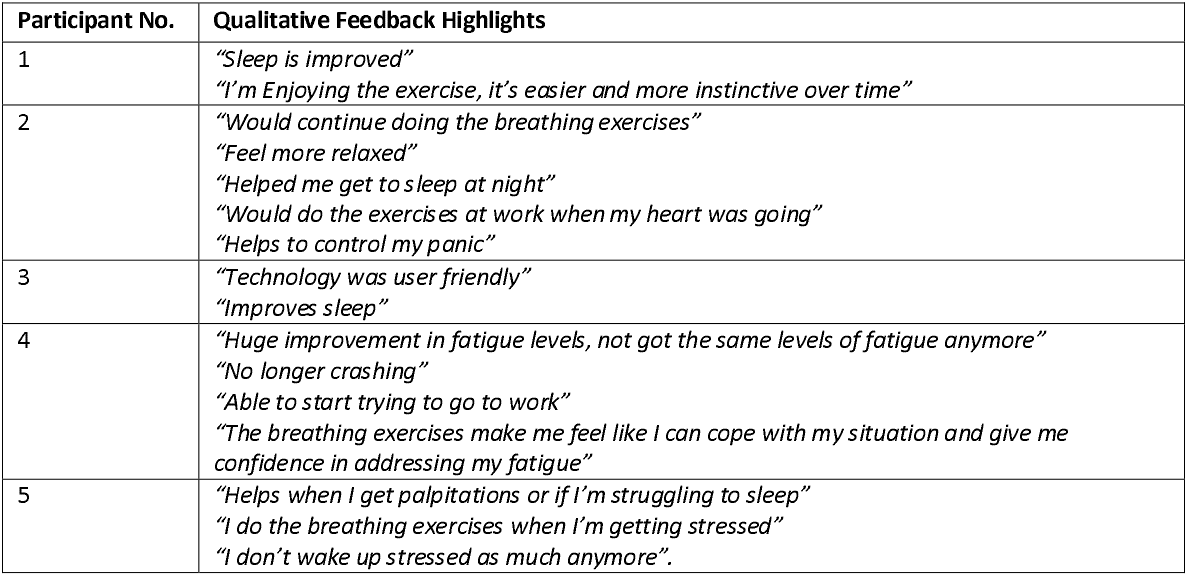
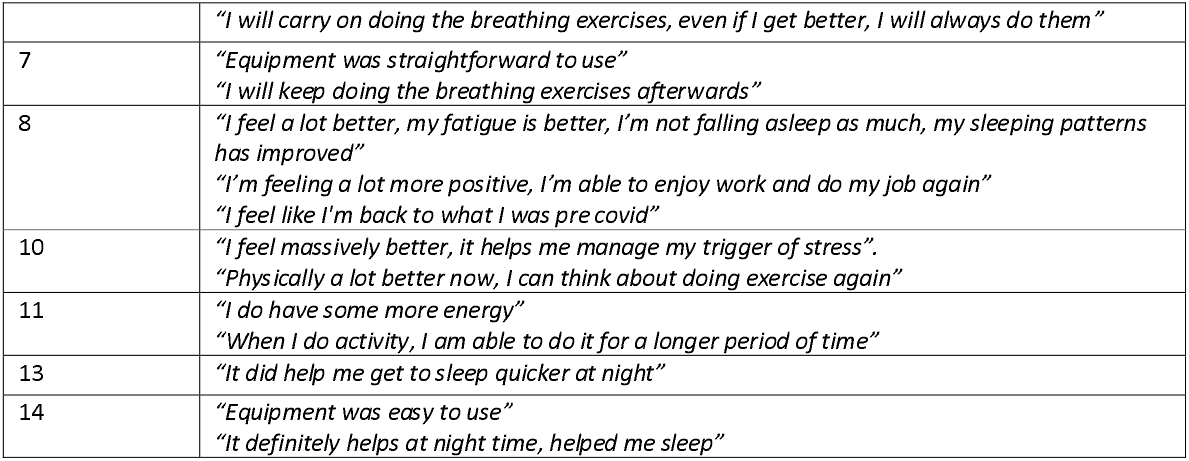
Participant feedback upon study completion.

## Discussion

This study demonstrated that an HRV-B intervention using diaphragmatic breathing, smartphone HRV application and an ECG chest strap was feasible for participants with LC in a home-based setting. This study represents the first use of HRV-B with diaphragmatic breathing modulation in a LC population. The study adds weight to the notion that dysautonomia plays a critical role in LC symptom generation and interventions to target this may prove beneficial. It should be noted that unlike many potential LC interventions, diaphragmatic breathing is a non-pharmacological treatment, well tolerated and scalable with app-based technologies. The results of this study support a further controlled trial in LC to explore its actual efficacy on symptoms, particularly dysautonomia.

Other studies of HRV-B as an adjunct in chronic disease management have found similar positive results amongst other conditions.^13^ In fibromyalgia, Hassett et al demonstrated significant improvement in fibromyalgia physical functioning scores which was evident at 3 months follow up (p-0.0022).^15^ As demonstrated by the qualitative feedback, participants were satisfied with the HRV-B practice which is comparable to other studies.^13^ There are many indices for capturing HRV and most significant improvement was the RMSSD metric, reflecting greater parasympathetic activity.^13^ This was also demonstrated by Shumann et al albeit in a different population of veterans with post-traumatic stress disorder, although no p-values were provided for comparison.^21^

All participants fed back that they appreciated the concept of the technology and particularly the NASA Lean Test^27^ and the adapted Autonomic Profile (aAP)^30^ assessment tools for dysautonomia. They found the tools easy to use and empowering. Other authors have noted that testing for dysautonomia should be more accessible and holistic.^10^ The aAP is both a holistic and comprehensive test assessing different triggers of dysautonomia including position, meal, time of day and exercise. This is the first clinical research study featuring its use and we would encourage its use in future research along with the NASA lean test for monitoring the condition and response to interventions.

HRV data collected in this study was diverse and rich. The Elite HRV data was collected only during the 10 minute breathing intervention. The Fitbit collected data during sleep offered a reasonable longer-term measure of HRV. There was a significant improvement in RMSSD from the Fitbit data, however the effect size was not as pronounced as clinical outcome measures. The reason for this may be due to the fact that HRV is impacted by many factors including nutrition, sleep, exercise and menstrual periods in females.^32-34^ We did not control for these factors and future trials should attempt to control for them to truly quantify effect of HRV-B on LC and dysautonomia symptoms.

The study had several limitations. It was a small open-label uncontrolled feasibility study, and we need to bear caution about the generalisability of study findings. The study has however showed that the technology can be used in a home setting independently by individuals with LC. The changes in physiological parameters did not match the clinical outcome measures in terms of significance values and effect sizes. This can be explained by the World Health Organisation’s International Classification of Functioning Disability and Health (ICF) framework which suggests non-linear changes in different domains of the health condition (body function, functional activities and quality of life). The intervention duration could be argued to be short to have a reasonable effect on autonomic imbalance and future research could explore the relationship between dose (duration) and response. Finally, we did not undertake a long follow-up after stopping the use of technology. This needs to be explored in future larger scale studies.

In summary, a diaphragmatic breathing technique using HRV-B is feasible to be used independently in a home setting by individuals with LC and the intervention seems to have a potential effect on improving LC symptoms, particularly those related to dysautonomia.

## Data Availability

All data produced in the present study are available upon reasonable request to the authors

## Ethics and dissemination

The study received ethical approval from Health Research Authority (HRA) Leicester South Research Ethics Committee (21/EM/0271).

## Acknowledgements

The authors would like to thank all study participants and the Leeds LC Advisory Group (PAG) for their involvement in all stages of this study.

## Patient and Public Involvement

Members of the PAG provided input to analyse and develop the research aims, objectives, and questions, ensuring these align with the key research priorities of those with LC and dysautonomia. All advisory group members have lived experience of LC. The PAG met quarterly to review progress, ensure the research continues to answer relevant issues and that findings can inform LC care.

## Author contributions

MS and AC conceptualised the study. MS, AC and RJOC were awarded EPSRC IAA pump-priming grant for the feasibility study with MS as the Principal Investigator. All authors contributed to the study design and obtained ethical approval. JC and NI performed the initial statistical analysis independently and verified the results. JC wrote an initial draft of the paper by adapting the grant proposal, the ethics protocol, and the protocol paper. All authors approved the final manuscript. MS is the corresponding author and guarantor.

